# Generalizability of Clinical Prediction Models in Mental Health - Real-World Validation of Machine Learning Models for Depressive Symptom Prediction

**DOI:** 10.1101/2024.04.04.24305250

**Authors:** Maike Richter, Daniel Emden, Ramona Leenings, Nils R. Winter, Rafael Mikolajczyk, Janka Massag, Esther Zwiky, Tiana Borgers, Ronny Redlich, Nikolaos Koutsouleris, Renata Falguera, Sharmili Edwin Thanarajah, Frank Padberg, Matthias A. Reinhard, Mitja D. Back, Nexhmedin Morina, Ulrike Buhlmann, Tilo Kircher, Udo Dannlowski, 2107 consortium, PRONIA consortium, MBB consortium, Tim Hahn, Nils Opel

**Affiliations:** Department of Psychiatry and Psychotherapy, Jena University Hospital, Jena, Germany; Institute for Translational Psychiatry, University of Münster, Münster, Germany; German Center for Mental Health (DZPG), Site Jena-Magdeburg-Halle, Germany; Center for Intervention and Research on adaptive and maladaptive brain Circuits underlying mental health (C-I-R-C), Jena-Magdeburg-Halle, Germany; Institute of Psychology, University of Münster, Münster, Germany; Joint Institute for Individualisation in a Changing Environment (JICE), University of Münster and Bielefeld University, Germany; Institute of Medical Epidemiology, Biometrics, and Informatics, Interdisciplinary Center for Health Sciences, Medical School of the Martin Luther University Halle-Wittenberg, Halle (Saale), Germany; Department of Psychology, Martin Luther University Halle-Wittenberg, Halle (Saale), Germany; Department of Psychiatry and Psychotherapy, University Hospital LMU Munich, Munich, Germany; Department for Psychiatry, Psychosomatic Medicine and Psychotherapy, University Hospital Frankfurt, Goethe University, Frankfurt am Main, Germany; Department of Psychiatry, University of Marburg, Marburg, Germany; Max Planck Institute for Metabolism Research, Cologne, Germany; German Center for Mental Health (DZPG), Site Munich-Augsburg, Germany; Department of Psychosis Studies, Institute of Psychiatry, Psychology and Neuroscience, King’s College London, United Kingdom; Max Planck Institute of Psychiatry, Munich, Germany

## Abstract

Mental health research faces the challenge of developing machine learning models for clinical decision support. Concerns about the generalizability of such models to real-world populations due to sampling effects and disparities in available data sources are rising. We examined whether harmonized, structured collection of clinical data and stringent measures against overfitting can facilitate the generalization of machine learning models for predicting depressive symptoms across diverse real-world inpatient and outpatient samples. Despite systematic differences between samples, a sparse machine learning model trained on clinical information exhibited strong generalization across diverse real-world samples. These findings highlight the crucial role of standardized routine data collection, grounded in unified ontologies, in the development of generalizable machine learning models in mental health.

**One-Sentence Summary:** Generalization of sparse machine learning models trained on clinical data is possible for depressive symptom prediction.

## Main Text

The inability to individually predict the occurrence of symptoms and their trajectories remains a major limitation for improving mental health care. Generating data-driven support for clinical decision-making and diagnostics is therefore the main objective of many innovations and advances in mental health research to date. To achieve this goal, we require machine learning models to learn consistent patterns for single participants from the complex and multi-faceted inter-individual variety present in real-world clinical populations, and for these models to be validated on independent datasets from a broad range of settings (*1*).

Systematic differences in available data sources and sampling effects between real-world clinical populations and those derived from research cohorts are thought to hinder generalizability of machine learning models in mental health (*2*–*4*). Clinical and demographic differences between and within research and real-world samples may lead to heterogeneity, which substantially impairs prediction accuracy and model generalizability (*5*). Assessing model generalization on real-world data is critical as they represent the populations for which predictions are intended, thus minimizing bias (*3*). While successful attempts have been made to train models for clinically relevant predictions within a single research dataset (*6*–*8*), previous investigations have often overlooked external validation, specifically validation in real-world clinical samples (*9*). Recently, attempts at validating models for treatment response prediction in mental health in unseen, independent data have failed, raising concerns about their generalizability (*10, 11*).

Given these recent concerns about the generalizability of models for clinical use cases such as treatment response prediction, it appears imperative to first determine whether robust and generalizable models for predicting the complex phenomena of mental health symptoms can indeed be achieved, especially considering the suspected heterogeneity across both research and real-world settings.

### Using clinical data for prediction

Although imaging and genetic data have proven to be invaluable for advancing precision medicine outside of mental health (*12*–*15*), previous mental health research has repeatedly demonstrated the particular relevance of training models on clinical information when predicting symptom trajectories and treatment outcome in disorders such as schizophrenia or depression (*16, 17*). However, despite the technical feasibility of implementing structured collection of clinical information, the widespread absence of harmonized machine-readable clinical data persists across research and clinical settings, primarily due to a lack of uniform data standards and shared ontologies in mental health.

If prediction using clinical data in easily implementable structured formats is not feasible, and if sampling biases or batch effects impede model generalizability to the extent that generalizable cross-sectional symptom prediction is not possible, then a reevaluation of our current direction is imperative. We therefore need to improve our understanding of the differences between study populations and real-world data and investigate the generalization of predictive models for mental health symptoms in unseen, independent data from various sites and settings as a foundation, before taking on the even more complex challenges of predicting symptom trajectories in response to intervention. Against this backdrop, the present study investigated whether structured clinical information facilitates the generalizability of a machine learning model for predicting depressive symptoms cross-sectionally across diverse samples, sites, and time points despite potential sampling and treatment effects. Specifically, we aimed to systematically validate a machine learning model trained on homogenous research data on real-world clinical data obtained from both inpatient and outpatient settings, as well as from the general population.

### Data sources

We evaluated sampling effects and model generalization across affective disorder patients from both a study population and a real-world sample recruited at the same psychiatric hospital: For the study sample (study population inpatients, site #1), we used clinical and self-report data from two pooled neuroimaging cohorts at the same site with virtually identical data assessment protocols. As comparison to the research setting, a sample from a naturalistic study of a real-world clinical population that was digitally phenotyped during inpatient treatment at the same psychiatric hospital was included (real-world inpatients, site #1). All available data were extracted and retained as predictor variables for the training of machine learning models if they were available in both samples. This resulted in a set of 76 features that were used to train a model for the prediction of depressive symptoms on study population inpatients #1 and tested on real-world population inpatients #1. Further information about all materials can be found in the supplementary material (SM, pp. 3-5).

To assess model generalization across different sites and settings, we included seven additional samples from various sites across Germany and one sample containing data from multiple sites across Europe, deviating further from the study population in terms of patient characteristics and recruitment setting with each site. To capture heterogeneity and diversity of real-world patient populations, these samples included inpatient samples with persistent depressive disorder (PDD) undergoing specialized psychotherapy, inpatient samples undergoing ECT treatment as well as outpatient samples from psychotherapy services undergoing long-term psychotherapeutic treatment, inpatient and outpatient participants with recent onset depression (ROD), and a general population sample with no relation to a clinical setting. An overview of all samples including descriptive and clinical information can be found in table 1. All samples are findable through the Meta-Data Study Repository of the German Centre for Mental Health (DZPG) (https://webszh.uk-halle.de/cohort-registry/).

**Table 1.** Overview and descriptive information for all sites.

| Sample | Recruitment site | Intervention | Treatment Duration in Days | Age | Gender | Baseline Depression | Depression FU | Extra-version | Neuroticism | GAF | Somatization | CTQ EA |
| --- | --- | --- | --- | --- | --- | --- | --- | --- | --- | --- | --- | --- |
|  |  |  | Mean (SD) | Range<br>Mean (SD) | m/f | Mean (SD) | Mean (SD) | Mean (SD) | Mean (SD) | Mean (SD) | Mean (SD) | Mean (SD) |
| **Study Population Samples** |  |  |  |  |  |  |  |  |  |  |  |  |
| **Study population inpatients, site #1 (n=366)** | Department of Psychiatry, University Hospital Münster, Germany | Medication, CBT | NA | 18-65<br>37.16<br>(12.86) | 158/208 | 39.82<br>(17.02) | NA | 39.08<br>(14.53) | 66.95<br>(15.50) | 54.43<br>(9.37) | 11.69<br>(7.79) | 11.18<br>(5.50) |
| **Study population in- & outpatients, site #1 (n=83)** | Department of Psychiatry, University Hospital Münster, Germany | Medication, ECT, CBT | 167.24<br>(108.71) | 19-66<br>33.72<br>(12.13) | 28/55 | 34.17<br>(18.81) | 26.29<br>(16.92) | NA | NA | 57.84<br>(12.63) | NA | 11.19<br>(5.57) |
| **Study population inpatients, site #2 (n=109)** | Department of Psychiatry, University of Marburg, Germany | Medication, CBT | NA | 18-63<br>36.75<br>(13.18) | 50/59 | 36.43<br>(16.39) | NA | 45.24<br>(16.13) | 66.03<br>(15.36) | 54.96<br>(8.89) | 14.05<br>(8.69) | 11.41<br>(5.25) |
| **Study population inpatients, site #3 (n=43)** | Department of Psychiatry and Psychotherapy, Jena University Hospital, Germany | Medication, CBT | NA | 18-67<br>39.23<br>(15.58) | 18/24 | 49.65<br>(20.07) | NA | 32.71<br>(10.58) | 56.04<br>(14.78) | 42.85<br>(11.35) | NA | 11.62<br>(5.74) |
| **Study population in- & outpatients, multisite (n=301)** | Ten international recruitment sites<sup>1</sup> | Medication, psychotherapy, counseling | NA | 15-41<br>25.4<br>(6.11) | 155/146 | 39.88<br>(19.37) | NA | 47.12<br>(16.19) | 64.14<br>(17.33) | 54.02<br>(12.28) | NA | 9.21<br>(4.22) |
| **Real-World Samples** |  |  |  |  |  |  |  |  |  |  |  |  |
| **Real-world inpatients, site #1 (n=352)** | Department of Psychiatry, University Hospital Münster, Germany | Medication ,CBT | 44.67 (23.23) | 18-81 39.3 (17.22) | 165/18 7 | 39.40 (18.05) | 21.38 (18.57) | 36.17 (18.78) | 68.60 (17.32) | 53.86 (9.18) | 12.22 (7.75) | 11.51 (5.79) |
| **Real-world inpatients, site #4 (n=161)** | Department of Psychiatry and Psychotherapy, Ludwig-Maximilian University Munich, Germany | Medication ,10-week CBASP | 70.98 (8.12) | 18-66 39.33 (12.55) | 68/93 | 48.95 (16.77) | 36.10 (21.41) | 33.81 (14.51) | 71.26 (13.77) | 46.42 (8.25) | NA | 14.40 (6.01) |
| **Real-world outpatients, site #5 (n=144)** | Psychotherapeutic Outpatient Unit, University of Halle, Germany | Medication ,CBT | 191.98 (12.25) | 19-60 28.76 (9.72) | 32/112 | 32.47 (17.32) | 19.07 (17.56) | NA | NA | 62.11 (11.68) | 8.07 (6.09) | 10.72 (4.76) |
| **Real-world outpatients, site #6 (n=252)** | Psychotherapeutic Outpatient Unit, University of Münster, Germany | Medication ,CBT | 613.91 (338.53) | 19-64 31.99 (11.38) | 105/14 7 | 37.84 (16.39) | 15.16 (14.35) | NA | NA | NA | 9.47 (7.56) | NA |
| **Real-world general population sample (n=1210)** | Institute of Medical Epidemiology, Medical Faculty of the Martin Luther University Halle-Wittenberg, Germany | NA | NA | 20-72 51.10 (11.08) | 367/84 3 | 25.63 (19.23) | NA | 50.42 (19.66) | 48.56 (19.13) | NA | NA | NA |

Note. <sup>1</sup> see supplementary material for details.
**Abbreviations.** CBASP = Cognitive Behavioral Analysis System of Psychotherapy, CTQ EA = Childhood Trauma Questionnaire, Emotional Abuse subscale, ECT = electro-convulsive therapy, GAF = Global Assessment of Functioning (37, 50), Depression = Percentage of maximum possible depression severity score, calculated from Beck Depression Inventory (35, 48) or Patient Health Questionnaire, depression scale (49), Depression FU = depression severity after treatment, Extraversion = Percentage of maximum possible extraversion score, calculated from Big Five Inventory-S (51), Big Five Inventory -2- S (43), NEO-Five Factor Inventory (42), Somatization = Symptom Checklist 90-Revised, somatization subscale

### Patients and outcomes

From May 2010 to February 2024, 2,808 participants aged 15 to 81 were included. All participants were diagnosed with major depressive disorder (MDD) and undergoing inpatient or outpatient treatment at the time of assessment, with the exception of the real-world general population sample, from which participants were selected who reported having received an MDD diagnosis at some point before the assessment. Symptomatic outcomes were assessed based on scores from self-report measures of depression severity for all sites (see SM, p. 5). Where available, depression severity after a psychotherapeutic intervention or at the conclusion of treatment was additionally included for model validation across time-points.

### Systematic Comparison between Study Populations and Real-World Samples

To systematically assess sample differences in clinical features and risk factors, we compared study population inpatients #1 and real-world inpatients #1, both consisting of participants recruited and treated at the same university hospital. Comparisons between the groups were assessed for the available variables, which could be grouped into the following dimensions: sociodemographic variables, current symptom severity, current psychotropic medication, family and personal psychiatric history, childhood maltreatment and stressful life events, somatic symptoms, and personality dimensions.

The two samples differed substantially in features from all dimensions except for somatic symptoms. The real-world sample displayed more severe current depressive symptoms only in external symptom assessment, not in a self-report measure. They also showed a more severe disease course, as well as differences in prescribed medication (more stimulants, benzodiazepines, and z-drugs), recalled childhood maltreatment (more physical neglect) and personality dimensions (lower extraversion and conscientiousness, higher agreeableness) compared to the study population (see SM, Table S2).

### Real-World Validation of machine learning model and development of sparse model

First, we trained a model on all N=366 study population inpatients #1, using all available 76 features to predict depression severity. Analogous to Chekroud et al. (*10*), we used the elastic net algorithm, a penalized regression method that is appropriate when covariates are correlated with one another and predictors may only be sparsely endorsed (see SM for more details; (*18, 19*). We performed cross-validation to assess validation performance of our model using the PHOTONAI software (www.photon-ai.com, (*20*)). The cross-validation part of this procedure randomly reshuffles the data and separates the dataset into 10 non-overlapping folds and uses 9 of the subsets for training, repeating the process such that each subset is left out once for testing. The repeated part of this procedure randomly reshuffles and re-splits the data ten times to reduce the impact of the first random data split; in aggregate, 100 total models were fit to the 10 folds by 10 repeats. Model performance was calculated by averaging the performance metrics across all 100 models. This procedure yielded an internal validation performance of Pearson r(364)=.57 (Standard Deviation = .151). Next, we identified the most relevant features for this model using permutation importance with 1,000 repeats. This yielded five main variables driving model performance (Figure 1): neuroticism, extraversion, global assessment of functioning, somatization, and emotional abuse during childhood, one of which (extraversion) had emerged as significantly different between study population and real-world inpatients in the previous analysis step. Using these five variables alone, we trained the base model on all N=366 study population inpatients #1. We then tested the base model based on the five relevant variables in N=352 real-world inpatients #1 consisting of participants recruited and treated at the same university hospital. Based on the prediction of the base model trained above, we computed the Pearson correlation between the true and the predicted values to assess predictive performance in the real-world sample. The base model performed above chance in the real-world sample (r(350)=.73, p<.001). Using the Binomial Effect Size Display (BESD, see SM) for illustration, this corresponds to an accuracy of 87% in a classification scenario.

**Fig. 1.**
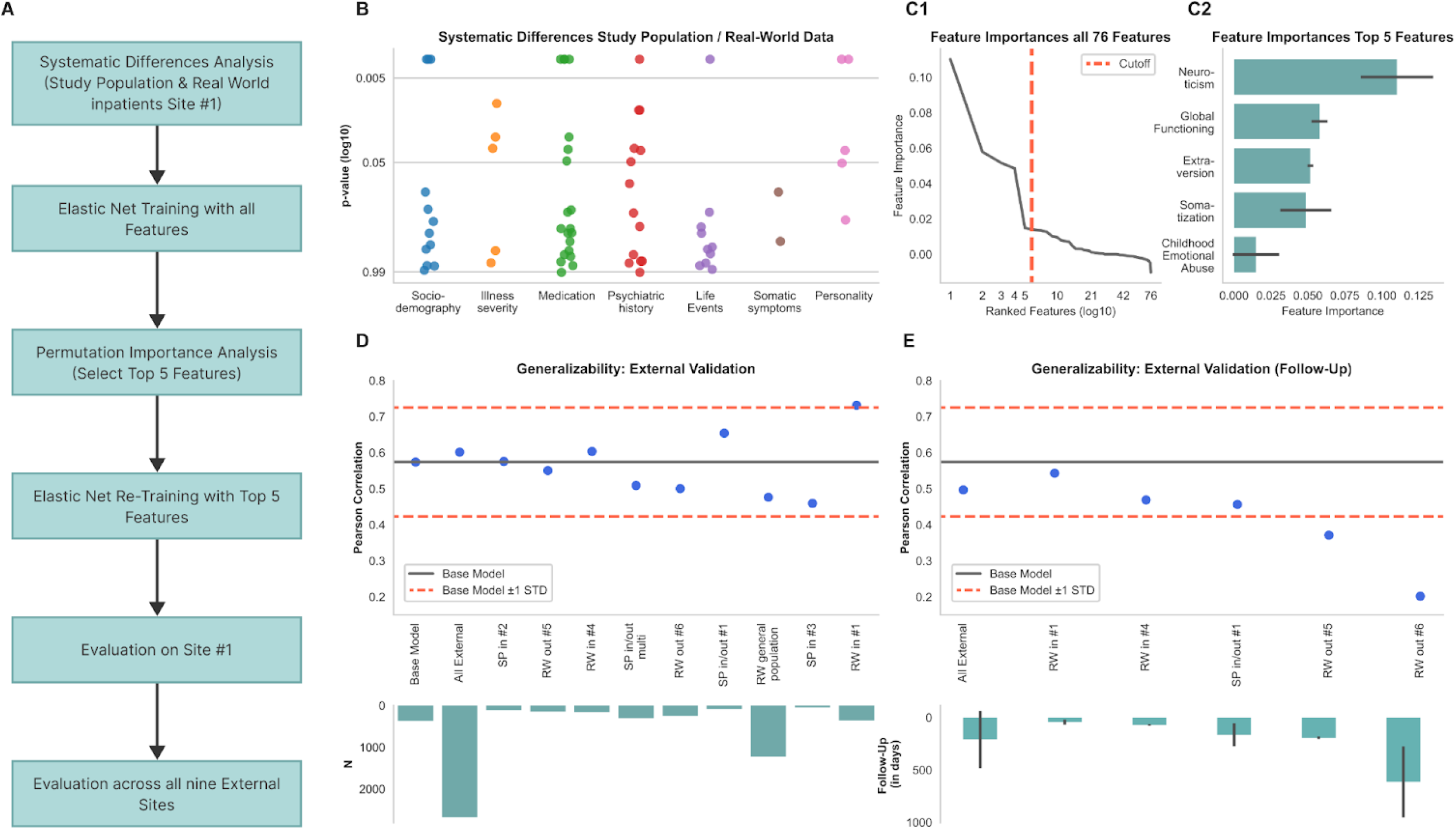
Methodology and results of the predictive model analysis. (**A**) Analytic workflow from systematic differences analysis to multisite model evaluation. (**B**) Scatter plot depicting p-values for group differences between study population and real-world inpatients from site #1 across clinical and demographic variables. (**C1**) Line plot of ranked feature importances with specified cutoff. (**C2**) 5 Bar plot highlighting the top 5 features selected through permutation importance analysis. (**D**) External validation results of the base model showing Pearson correlation of true and predicted depressive symptoms, contrasted across nine external sites. (**E**) Follow-up validation scatter plot showing Pearson correlation of true and predicted depressive symptoms following therapeutic intervention, including the presentation of average follow-up durations by site.

### Generalizability of the base model across sites, treatment settings, and populations

To further assess model generalizability, we tested the base model across all nine external samples from different research and clinical settings and geographical sites with a total of N=2,675 participants for external validation (see Figure 1). The model performed above chance level across all external datasets (r(2,673)=.60, Standard Deviation = .089, p<.001). Using the BESD for illustration, this corresponds to an accuracy of 80% in a classification scenario. Importantly, this performance is nominally higher than base model performance on study population inpatients #1 dataset (see above), indicating excellent generalization performance.

Investigating performance on the nine samples separately shows that performance on all sites varies between r(1,227)=.48 in the real-world general population sample, r(250)=.50 in real-world outpatients #6 and r(350)=.73 in real-world inpatients #1. Thus, even the lowest performance (real-world general population sample) lies within .60 standard deviations of the mean of the base model performance (r(364)=.57, standard deviation=.151). Note that the comparatively poorer performance in the real-world general population sample may result from only two of the five features being available for this sample, which moreover differed most markedly from the training set in participant characteristics due to it being a general population sample in which participants were not necessarily acutely depressed or currently undergoing treatment. Supplementary analyses excluding the most highly weighted feature, neuroticism, also confirmed good generalizability of the sparse model across sites (see SM, p. 9).

### Generalizability of machine learning model across two time points

To assess whether base model performance remains robust after therapeutic interventions, we used the base model to predict depression severity after treatment. We show that the base model performs above chance level (r(566)=.50, p<.001) across the five external datasets which provide an assessment after a therapeutic intervention (study population in-& outpatients #1, real-world inpatients #1, real-world inpatients #4, real-world outpatients #5, real-world outpatients #6). Again, using the BESD for illustration, this corresponds to an accuracy of 75% in a classification scenario. While this performance is nominally lower than base model performance on the study population inpatients #1 dataset, it lies within one standard deviation of the mean of the base model performance, indicating good generalization for the prediction of depression severity at a different measurement time without explicit training. Investigating performance on the five sites separately shows that performance varies between r(125)=.20 (real-world outpatients #6) and r(56)=.54 (real-world inpatients #1). Note that treatment duration differed substantially between sites and **treatment modalities. The comparatively low performance in real**-world outpatients #6 may be due to the long duration of treatment. Investigating this, we show that treatment duration is indeed positively associated with model error across all sites indicating increased model error with longer duration between baseline and follow-up assessment (Spearman r(554)=0.12, p=0.004). Investigating potential model bias, we assess the association of model error and age and sex, respectively. We show that neither age (Spearman r(554)=0.07, p=0.093 nor sex (t(554)=-1.54, p=0.123) are significantly associated with model error. Supplementary analyses indicate a classification accuracy of 66% for identifying subjects with persistent depressive symptoms at both time points based on the top 5 variables (see SM, p. 8).

## Discussion

In this study, we demonstrate that a machine learning model trained on mental health research data can achieve comparable performance for predicting depression severity in unseen, independent real-world datasets across different sites, treatment settings, and time points. To the best of our knowledge, this study includes the most extensive independent validation in the field of mental health research to date. In contrast to previous studies (*10, 16*), we show robust generalization performance across nine independent sites comprising over 2,600 participants, reflecting the full spectrum of heterogeneity and diversity present in real-world patient populations. This suggests that real-world validation of mental health symptom prediction models is possible, despite substantial sample heterogeneity.

### Tackling the challenges of model generalization

A first challenge to consider for model generalization is the avoidance of overfitting when training the base machine learning model (*21*). When a model overfits, it captures both the signal and the noise in the training data on which it may perform exceptionally well while failing to generalize to new, unseen data (*22*). Regularization, which imposes constraints on the model parameters to encourage sparsity, can help prevent overfitting by promoting simpler, more interpretable models. In our study, working with low-dimensional clinical data and further reducing the dimensionality of the feature space by focusing on the most informative features was used to prevent overfitting.

### Sampling effects between study and real-world populations

The second challenge for validating models in independent datasets is that patient groups from research contexts may be too different from real-world clinical populations (*21*). We demonstrate that systematic differences indeed exist between research populations and real-world MDD patients, even when both samples are treated and assessed at the same psychiatric hospital. However, we also demonstrate that these differences do not necessarily impede model generalization to populations from different study sites or real-world treatment contexts. While previous research from other areas of medicine, such as predicting positive COVID-19 screenings, reveal that site-specific model customization can improve predictive performance, the approach of applying a ready-made model “as-is” has been found to be effective (*23*) and appears to also be feasible in the context of mental health.

### Predicting symptom severity at different time points

Additionally, biases arise not only from baseline differences in patient characteristics and site but also from variations in treatment modalities, especially for prospective predictions of depression severity after a mental health intervention. We show that our model remains robust after treatment with markedly different modalities and across various settings, particularly for the translation from inpatient to outpatient psychotherapy service users. While performance drops markedly the further the treatment context deviates from the training set and with increasing time between baseline and follow-up assessment, prediction of both baseline as well as post-treatment depression severity is still possible. This underlines the finding that heterogeneity within and between datasets and measurement time does not stand in the way of model generalizability. Although the predictive clinical features used in our sparse model may allow for the identification of participants with persistent depressive symptoms across time points and after treatment, it should not be misinterpreted as a readily applicable model for clinical decision support. The present findings rather suggest the general feasibility of developing machine learning models for predicting complex phenomena of mental health symptoms. These findings may thus serve as a foundational step for future endeavors aimed at refining models suitable for ecologically valid clinical use cases in daily practice.

### Predictive value of clinical information

Another challenge of model generalization is the quality, quantity, and diversity of the data needed to achieve accurate predictions. While previous research in study populations shows that predictive models which include more than one data modality, such as clinical, neuroimaging, and genetic data, achieve better performance (*24*) we demonstrate that symptom severity prediction is possible with sparse features that can be collected during the clinical routine. This is in line with previous findings on the particular importance of clinical information when predicting symptom trajectories and treatment outcome in mental health research (*16, 17*). The extracted features, encompassing two personality dimensions, somatic symptom severity, childhood emotional abuse, and global functioning, and thus a mixture of state and trait variables, consistently form a predictive pattern for depression severity across diverse patient populations, irrespective of illness stage or treatment setting. It is crucial to highlight that these features have demonstrated greater importance compared to more than 70 other variables, some of which might be presumed to hold equal or greater relevance in determining depressive symptom severity including clinician-relevant factors like psychiatric history or prescribed medication. However, it is crucial to note that the initial selection of 76 features may not encompass the full spectrum of variables with predictive potential and that there may be other variables of greater significance.

### Improving structured clinical data collection

Given our demonstration of the generalizability of machine learning models trained on clinical information, along with considerations of technical and cost efficiency, these findings should encourage structured, machine-readable clinical information acquisition in routine settings. We should thus increase efforts to improve interoperability and invest in uniform data standards and ontologies in mental health. Successful examples from the medical community such as the introduction of the Systematized Nomenclature of Medicine, Clinical Terms (SNOMED CT (*25*)), Logical Observation Identifiers, Names, and Codes (LOINC (*26*)), and Fast Health Interoperability Resources (FHIR (*27*)) profiles are encouraging in this regard. Wide-reaching infrastructures such as the German Medical Informatics Initiative (*28*) as well as other international efforts (*29*–*31*) have set the goal of improving integration of clinical data from patient care and medical research and the French Health Data Hub is even explicitly set up to facilitate health data sharing with the aim of developing health-related Artificial Intelligence projects (*32*). As dedicated solutions for mental health are lacking within current infrastructure efforts, our findings highlight the necessity for national and international endeavors to tailor, develop, and disseminate such solutions specifically for mental health. The recent establishment of the German Centre for Mental Health (DZPG) with its translational agenda and integration with key data infrastructures in Germany signifies an important step forward in this regard (*33*).

In summary, our findings highlight successful real-world validation of sparse machine learning models for depressive symptom prediction and emphasize the potential of using standardized routine data collection for developing generalizable empirical models in mental health.

## Data Availability

All samples and their data are findable and requestable through the Meta-Data Study Repository of the German Centre for Mental Health (DZPG). The machine learning model will be published in the PHOTONAI model repository.

https://webszh.uk-halle.de/cohort-registry/

## Acknowledgments

We are deeply indebted to all participants in this study.

## Funding

Interdisciplinary Center for Clinical Research (IZKF) of the medical faculty of Münster grant SEED 11/18 (NO), ), Dan3/022/22 (UD)

German Research Foundation grants RE4458/1-1 (RR), KI 588/14-1 (TK), KI 588/14-2 (TK), KI 588/15-1 (TK), KI 588/17-1 (TK), DA 1151/5-1 (UD), DA 1151/ 5-2 (UD), DA 1151/6-1 (UD), DA1151/9-1 (UD), DA1151/10-1 (UD), DA1151/11-1 (UD), KR 3822/5-1 (AK), KR 3822/7-2 (AK), NE 2254/1-2 (IN), NE 2254/2-1 (IN), NE2254/3-1 (IN), NE2254/4-1 (IN), HA 7070/2-2 (TH), HA7070/3 (TH), HA7070/4 (TH), KO-121806 (KD), and JO22022/1-1

Collaborative Project funded by the European Union (EU) under the 7th Framework Programme grant 601252

German Federal Ministry of Education and Research grants 01EE2305C (RR), 01EE230A (NO), 01EE2303A, 01ER1301A/B/C, 01ER1511D, 01ER1801A/B/C/D, the Federal States of Germany and the Helmholtz Association, the participating universities and the institutes of the Leibniz Association

FöFoLePLUS program of the Faculty of Medicine of the Ludwig-Maximilians-University, Munich, Germany, grant #003, MCSP (MAR)

## Author contributions

Conceptualization: NO, MR, DE, TH

Data curation: MR, RF, SET, JM

Methodology: NO, TH, DE, MR

Formal analysis: MR, DE, TH, RF

Funding acquisition: UD, NO, TK, RR, NK

Project administration: MR

Supervision: NO

Writing – original draft: MR, DE, NO, TH

Writing – review & editing: MR, DE, RL, NRW, RM, JM, EZ, TB, RR, NK, RF, SET, FP, MAR, MDB, NM, UB, TK, UD, TH, NO, PB, RU, FF, RKRS, JK, SB, EML, AB, RL, SM, KT, KF, NS, AK, JG, IN, BS, NA, HJ, AJ, FS, KB, FTO, PU, LT, JR, RB, LFK, MR

## Competing interests

FP is a member of the European Scientific Advisory Board of Brainsway Inc., Jerusalem, Israel, and the International Scientific Advisory Board of Sooma, Helsinki, Finland. He has received speaker’s honoraria from Mag&More GmbH and the neuroCare Group. His lab has received support with equipment from neuroConn GmbH, Ilmenau, Germany, and Mag&More GmbH and Brainsway Inc., Jerusalem, Israel.

MAR has received financial research support from the EU (H2020 No. 754740 ) and served as PI in clinical trials from Abide Therapeutics, Böhringer-Ingelheim, Emalex Biosciences, Lundbeck GmbH, Nuvelution TS Pharma Inc., Oryzon, Otsuka Pharmaceuticals and Therapix Biosciences.

The remaining authors declare that the research was conducted in the absence of any commercial or financial relationships that could be construed as a potential conflict of interest.

## Data and materials availability

All samples and their data are findable and requestable through the Meta-Data Study Repository of the German Centre for Mental Health (DZPG) (https://webszh.uk-halle.de/cohort-registry/). The machine learning model will be published in the PHOTONAI model repository.

## Supplementary Materials

List of Group Authors

Materials

Methods

Figs. S1 and S2

Tables S1 and S2

References (*35-59*)

## Notes

### Funding Statement

Interdisciplinary Center for Clinical Research (IZKF) of the medical faculty of Muenster grant SEED 11/18 (NO), ), Dan3/022/22 (UD)
German Research Foundation grants RE4458/1-1 (RR), KI 588/14-1 (TK), KI 588/14-2 (TK), KI 588/15-1 (TK), KI 588/17-1 (TK), DA 1151/5-1 (UD), DA 1151/ 5-2 (UD), DA 1151/6-1 (UD), DA1151/9-1 (UD), DA1151/10-1 (UD), DA1151/11-1 (UD), KR 3822/5-1 (AK), KR 3822/7-2 (AK), NE 2254/1-2 (IN), NE 2254/2-1 (IN), NE2254/3-1 (IN), NE2254/4-1 (IN), HA 7070/2-2 (TH), HA7070/3 (TH), HA7070/4 (TH), KO-121806 (KD), and JO22022/1-1
Collaborative Project funded by the European Union (EU) under the 7th Framework Programme grant 601252
German Federal Ministry of Education and Research grants 01EE2305C (RR), 01EE230A (NO), 01EE2303A, 01ER1301A/B/C, 01ER1511D, 01ER1801A/B/C/D, the Federal States of Germany and the Helmholtz Association, the participating universities and the institutes of the Leibniz Association
FoeFoLePLUS program of the Faculty of Medicine of the Ludwig-Maximilians-University, Munich, Germany, grant #003, MCSP (MAR)

### Author Declarations

Ethics committee of the medical association Westphalia-Lippe and the University of Muenster, officially entitled "Ethikkommission der Aerztekammer Westfalen-Lippe und der Westfaelischen Wilhelms-Universitaet Muenster", gave ethical approval for the entire study.

