## Supplementary Materials for "Generalizability of Clinical Prediction Models in Mental Health - Real-World Validation of Machine Learning Models for Depressive Symptom Prediction"

**The PDF file includes:**

List of Group Authors

Materials and Methods
Figs. S1 and S2
Tables S1 and S2
References (*35-59*)

List of Group authors

*PRONIA consortium*

Department of Pathophysiology and Transplantation, University of Milan: Paolo Brambilla

Institute for Mental Health and Centre for Human Brain Health, School of Psychology, University of Birmingham: Rachel Upthegrove

Department of Psychiatry, University of Udine: Franco Fabbro

Department of Psychiatry, University of Turku: Raimo K. R. Salokangas

Department of Psychiatry and Psychotherapy, Faculty of Medicine and University Hospital, University of Cologne: Joseph Kambeitz

Department of Psychiatry, Psychiatric University Hospital, University of Basel: Stefan Borgwardt

Department of Psychiatry and Psychotherapy, Medical Faculty, Heinrich-Heine University, Düsseldorf: Eva Meisenzahl-Lechner

Department of Basic Medical Science, Neuroscience, and Sense Organs, University of Bari Aldo Moro: Alessandro Bertolino

Department of Psychiatry and Psychotherapy, University of Münster: Rebekka Lencer

*FOR2107 consortium*

Institute for Translational Psychiatry, Münster, Germany: Susanne Meinert, Katharina Thiel, Kira Flinkenflügel, Navid Schürmeyer, Anna Kraus, Janik Goltermann

Department of Psychiatry, University of Marburg, Germany: Igor Nenadic, Benjamin Straube, Nina Alexander, Hamidreza Jamalabadi, Andreas Jansen, Frederike Stein, Katharina Brosch, Florian Thomas-Odenthal, Paula Usemann, Lea Teutenberg

*MBB consortium*

Department of Psychiatry and Psychotherapy, Jena University Hospital, Jena, Germany: Janette Ratzsch, Rogério Blitz, Lena Florentine Köhler, Moritz Rau

Materials – Analysis Step 1

For the group comparison between study population and real-world inpatients from site #1, available data were grouped along the following eight dimensions:

***Sociodemographic variables***

In addition to age and gender, sociodemographic variables encompassed the number of children and siblings, the highest education degree of the participants and both their parents and the average monthly household income. It was also recorded whether participants or their parents immigrated to Germany. The population size of the main place of residence during childhood and adolescence was also recorded. From the population size, an urbanicity score was calculated in which a score of 1 was assigned to rural areas with a population size below 10000, 2 – towns with a population size between 10000 and 100000, and 3 – cities with a population of over 100000 (*34*).

***Depressive symptom severity and level of functioning***

The current severity of depressive symptoms was assessed with Beck Depression Inventory (BDI) (*35*)) as a self-report questionnaire with 21 items. The 17-item Hamilton Depression Scale (HAMD) (*36*) was carried out by trained raters as an objective measure of depression severity. Within this dimension, percentage of diagnosed single depressive episodes vs recurring depression and percentage of participants with psychiatric comorbidities were also investigated. Global Assessment of Functioning (GAF) (*37*) was used by trained raters in both samples to assess participants’ psychological, social, and occupational functioning on a scale from 1 (severely impaired) to 100 (extremely high functioning).

***Current psychotropic medication***

Type and dose of psychopharmacological treatment were recorded by the research team in study population inpatients and extracted from the electronic health record (EHR) in real-world inpatients #1. A strategy described by Redlich et al. (2014) was subsequently applied: Each psychotropic medication was coded as absent (0), low (1; defined as equal or lower average dose), or high (2; defined as greater than average dose), relative to the midpoint of the daily dose range recommended by the Prescribers’ Digital Reference (*38*). A composite measure of total medication load was calculated, reflecting dose and variety of different medications taken, by summing all individual medication scores. Moreover, each drug was assigned to its medication class and the prescription rates of each medication class between the samples were compared.

***Family and own psychiatric history***

Patients provided a detailed psychiatric history, including number and duration of previous inpatient treatments, their age at the time they were first treated due to psychiatric symptoms and the duration they had spent on sick leave or in retirement due to mental illness. In study population inpatients, the main diagnosis as well as psychiatric comorbidities were verified via SCID-I performed by trained raters, whereas in real-world inpatients, all diagnoses were directly entered into the EHR by the treating physician after admission and initial clinical assessment.

To assess family history and genetic risk for mental disorders, patients were asked whether an immediate family member had been formally diagnosed with a mental illness and to choose the disorder from a list including MDD, bipolar disorder (BD), substance use disorders, anxiety disorders, post-traumatic stress disorder (PTSD), obsessive-compulsive disorder (OCD), psychotic disorders, attention-deficit (and hyperactivity) disorders (ADHD), eating disorders, or other.

***Childhood maltreatment and stressful life events***

The Childhood Trauma Questionnaire (CTQ) (*39*) was used as a retrospective self-report measure for childhood maltreatment. It contains 28 items that are scored on a five-point Likert scale. The five subscales of childhood maltreatment that are measured by this instrument are physical neglect, emotional neglect, and physical, emotional, and sexual abuse. As a measure of more recent stressors, the Life Events Questionnaire (*40*) was used in study population inpatients to assess whether participants had encountered stressful events (a separation or divorce, death of a loved one, losing their job, and moving house) in the last six months before participation and the impact (from – 3, “very negative” to + 3, “very positive”) this had on respondents. In real-world inpatients, a modified version of the questionnaire was used for brevity, in which the impact was not assessed. The groups were therefore only compared on the frequency of encountering stressful life events and not on their impact.

***Somatic symptoms***

Somatic symptoms in the last 4 weeks before study inclusion were assessed for both samples based on the somatization subscale of the Symptom Check List (SCL-90-R) (*41*) which consists of 12 items describing somatic complaints. Participants rated each item on a 5-point Likert scale (0, “no problem” to 4, “very serious”). Body mass index (BMI) was calculated from height and weight which study population inpatients self-reported whereas, in real-world inpatients, these measures were extracted from the EHR for those participants for whom they were available from clinical routine documentation. For real-world inpatients, somatic comorbidities as diagnosed by the treating physician were extracted from the EHR and participants also indicated their somatic ailments on a list of somatic disorder categories. In study population inpatients, an extensive list of somatic disorders was applied as exclusion criteria before initial participation. This information can be found in table S1.

***Big Five Personality Dimensions***

Big Five personality dimensions were assessed with self-report questionnaires. In study population inpatients, the 60-item NEO-FFI (*42*) was used while real-world inpatients completed the 30-item BFI-2-S (*43*). Although these questionnaires were both constructed to measure the same underlying constructs (*44*), they are not directly comparable due to diverging scale ranges. To achieve comparability, a procedure described by Fleeson & Gallagher (2009) (*45*) was used to transform NEO-FFI and BFI-2-S values into absolute percent of maximum possible (POMP) scores (*46*). The ‘pomp’ function from the ‘fancyr’ package in R was used for score transformation (*47*). The samples were subsequently compared along their Big Five dimensions using these scores.

Materials – Analysis Step 2

All variables described above (with the exception of HAMD, which also measures depression severity) were included to train the first model in study population inpatients #1. This yielded five main features driving model performance. Using these five features alone, we trained the base model on all study population inpatients #1. We tested the base model based on the five features at external samples from different research and clinical settings and geographical sites. To make the variables comparable across different sites, some harmonization was conducted as outlined below:

***Outcome***

As depression severity, which was our outcome of interest, was assessed with different instruments (BDI (*35*), BDI-II (*48*), or PHQ-9 (*49*)) across different sites, we harmonized these measures by transforming them into absolute percent of maximum possible (POMP) scores as described above. The score represents the percentage a participant achieved in relation to the maximum possible depression severity that can be achieved in the measurement tool (*46*).

***Five Top Features***

***Global functioning***

As described above, GAF (*37*) was evaluated by trained raters to assess participants’ psychological, social, and occupational functioning on a scale from 1 (severely impaired) to 100 (extremely high functioning). In the study population in-& outpatient multisite sample, the split version of the GAF (*50*) was used which divides the scale into disability and symptomatic functioning. The mean of the two values was calculated to harmonize this variable with the single score available in the other sites.

***Childhood maltreatment***

The emotional abuse subscale of the CTQ (*39*) described above was included in the sparse model. No harmonization between sites was necessary.

***Somatic symptoms***

The somatization subscale of the SCL-90-R (*41*) as described above was included in the sparse model. No harmonization between sites was necessary.

***Big Five Personality dimensions***

Neuroticism and extraversion both emerged as features with most predictive value for the sparse model. As Big Five personality dimensions were assessed with different instruments between the sites (NEO-FFI (*42*), BFI-2-S (*43*), BFI-S (*51*)), they were transformed into POMP scores as described above, to allow for inter-site comparability (*45*, *46*).

Methods

Recruitment procedures and site information

*Study population inpatients, site #1*

For the study population inpatients, site #1, data sets from two independent cohorts were combined: the Marburg–Münster-Affective-Disorders-Cohort-Study (MACS - Münster site; (*52*)) and the Münster-Neuroimaging-Cohort (MNC; (*53*)). From both ongoing studies that comprise participants between 18–65 years of age, those with a confirmed MDD diagnosis during an acute depressive episode according to the German version of the Structured Clinical Interview for DSM-IV (SCID; (*54*)), who were currently undergoing inpatient treatment at the Department of Psychiatry, University Hospital Münster were selected. Any history of neurological (e.g., concussion, stroke, tumor, neuroinflammatory diseases) and medical (e.g., cancer, chronic inflammatory or autoimmune diseases, heart diseases, diabetes mellitus, infections) conditions were exclusion criteria. Patients received inpatient treatment as usual, including psychiatric medication and CBT-based psychotherapy.

*Real-World inpatients, site #1*

The real-world inpatients, site #1 sample consisted of psychiatric inpatients, aged 18 years or older, at the Department of Psychiatry, University Hospital Münster. Criteria for initial eligibility were the admission to any of the open adult inpatient services of the hospital, treatment duration of more than three days and the diagnosis of major depressive disorder (MDD) at the time of admission. Patients received inpatient treatment as usual, including psychiatric medication, CBT-based psychotherapy, electro-convulsive therapy (ECT), and repetitive transcranial magnetic stimulation (rTMS).

*Study population in-& outpatients, site #1*

The study population in- & outpatients, site #1 sample comprises participants between 18–65 years of age, those with a confirmed MDD diagnosis during an acute depressive episode according to the German version of the SCID, who were currently undergoing inpatient treatment at the Department of Psychiatry, University Hospital Münster or outpatient treatment at the psychotherapeutic outpatient unit, University Münster. Any history of neurological (e.g., concussion, stroke, tumor, neuroinflammatory diseases) and medical (e.g., cancer, chronic inflammatory or autoimmune diseases, heart diseases, diabetes mellitus, infections) conditions were exclusion criteria. Patients received inpatient treatment including psychotherapy, medication, and ECT, or outpatient CBT-based psychotherapy and medication and were assessed again at the conclusion of their treatment.

*Study population inpatients, site #2*

For the study population inpatients, site #2 sample, the Marburg site of the MACS cohort study was used. Participant selection and recruitment procedures were identical to study population inpatients at site #1, but all patients were treated and recruited at the Department of Psychiatry and Psychotherapy, University of Marburg. Patients received inpatient treatment as usual, including psychiatric medication and CBT-based psychotherapy.

*Study population inpatients, site #3*

For study population inpatients, site #3, participant selection and recruitment procedures were again harmonized to procedures from study population inpatients at sites #1 and #2, but all patients were treated and recruited at the Department of Psychiatry and Psychotherapy, University Hospital Jena. Patients received inpatient treatment as usual, including psychiatric medication, CBT-based psychotherapy, ECT, and rTMS.

*Study population in- & outpatients, multisite*

For the study population in- & outpatient multisite sample, participants were recruited at ten international sites as part of the Personalized Prognostic Tools for Early Psychosis Management (PRONIA; https://www.pronia.eu/) study, following a standardized recruitment and ascertainment protocol (see (*17*)). The recruitment sites included the Department of Psychiatry and Psychotherapy, Ludwig-Maximilians-University Munich, Germany; Department of Psychiatry and Psychotherapy, University of Cologne, Germany; Heinrich Heine University Düsseldorf, Germany; University of Münster, Germany; Department of Pathophysiology and Transplantation, University of Milan, Italy; University of Bari Aldo Moro, Italy;  University of Birmingham, United Kingdom; Department of Psychiatry, University of Udine, Italy; Department of Psychiatry, University of Turku, Finland; Department of Psychiatry and Psychotherapy, University of Basel, Switzerland. General inclusion criteria of the PRONIA study were age between 15 and 40 years, sufficient language skills for participation as well as capacity to provide informed consent/assent. General exclusion criteria were an IQ below 70, current or past head trauma with loss of consciousness (> 5 minutes), current or past known neurological or somatic disorders potentially affecting the structure or functioning of the brain, current or past alcohol dependence, or polysubstance dependence within the past six months, and any medical indication against MRI. Participants were selected for this sample if they met criteria for recent onset depression, meaning a diagnosis of MDD within the past 3 months, as established by SCID. Specific exclusion criteria for this sample were: (1) a previous episode of DSM-IV-TR major depression prior to the current or recent episode, and (2) a duration of the current episode exceeding 24 months. Participants received treatment as usual, including psychiatric medication, inpatient or outpatient psychotherapy, and counseling sessions.

*Real-world  inpatients, site #4*

The real-world inpatients sample #4 comprises participants who were recruited at the Department of Psychiatry and Psychotherapy of the LMU University Hospital, LMU Munich (German Clinical Trial Register ID: DRKS00019821). Patients between the age of 18 to 65 who were admitted to a structured 10-week inpatient psychotherapy program of Cognitive-Behavioral-Analysis System of Psychotherapy (CBASP; (*55*)) were eligible to take part in the ongoing naturalistic study. This program is primarily indicated for patients with chronic depression and general CBASP eligibility was assessed in a preliminary medical interview. Additionally, the main diagnosis of a persistent depressive disorder (PDD) was verified with SCID after admission. Patients with acute suicidality, unstable treatable somatic illnesses, such as acute or chronic infections, a current pregnancy or patients admitted to a second or subsequent CBASP booster session, were excluded. Baseline measures took place within the first 10 days while post measures were taken at the earliest 10 days before the planned discharge after 10 weeks.

*Real-world  outpatients, site #5*

The real-world outpatients, site #5 sample comprises participants with a confirmed MDD diagnosis during an acute depressive episode according SCID, aged 18 years or older, who were currently undergoing outpatient treatment at the psychotherapeutic outpatient unit, University of Halle. Criteria for initial eligibility were the admission of adult outpatient services, a diagnosis of major depressive disorder (MDD) at the time of admission, and an outpatient treatment for at least 6 weeks. All patients received CBT-based psychotherapy.

*Real-world  outpatients, site #6*

The real-world outpatients, site #6 sample includes participants with a confirmed MDD diagnosis according to SCID, who were currently undergoing CBT-based outpatient treatment at the psychotherapeutic outpatient unit, University Münster. All participants who were additionally recruited to take part in the study population in/ & outpatients, site #1 study during their treatment were excluded from this sample to make sure the samples were independent.

*Real-world general population sample*

For the real-world general population sample, participants from the general population 20-69 years old were invited to the study center, where an extensive program including interview, touch screen and various examinations took place. For a detailed description see (*56*). From all 10 000 participants at the Halle (Saale) site, those who self-reported having received a formal MDD diagnosis at some point in their lives were selected as the real-world general population sample.

Group statistics

For group comparisons, statistics were computed using IBM SPSS Version 26. For all comparisons, Benjamini-Hochberg false discovery rate (FDR)-corrected p-values were generated. Group comparisons were calculated with independent two-sample t-tests for continuous outcome variables, Χ²-tests for dichotomous outcome variables and Mann-Whitney-U test as a nonparametric test for ordinal outcome variables.

Machine Learning

We used the elastic net algorithm, a penalized regression method that is appropriate when covariates are correlated with one another and predictors may only be sparsely endorsed. We computed 10-fold cross-validation using 10 repeats to assess validation performance of our model using the PHOTONAI software ([www.photon-ai.com](http://www.photon-ai.com), (*20*)). The cross-validation part of this procedure separates the data set into 10 random folds and uses 9 of the subsets for training, repeating the process such that each subset is left out once for testing. The repeated part of this procedure re-splits the data ten times to reduce the impact of the random data split; in aggregate, 100 total models were fit to the 10 folds by 10 repeats. Model performance was calculated by averaging the performance metric across all 100 models. Based on the prediction of this baseline model, we computed Pearson correlations between the true and the predicted values to assess predictive performance.

For illustration, we computed the Binomial Effect Size Display (BESD (*57*)) from the Pearson correlations. In short, the BESD provides an approximation of the proportion of correct guesses about the direction of a correlation. It adjusts the initial chance level (50%) by incorporating the strength of the correlation coefficient, which represents the magnitude and direction of the relationship between two variables. The BESD offers a simple yet informative metric for evaluating the practical significance of correlation coefficients.

Classification of severely depressed non-responders

As the predictive model showed robust performance for depressive symptom prediction at two distinct time points before and after intervention, we additionally aimed to assess whether the same variables could be used to train a model to identify subjects with severe depressive symptoms at both time points thus allowing to assess its potential value for individual risk assessment. We used the established BDI cut-off of 29, indicating severe depression, which corresponded to a POMP score of 46.03 to stratify the sample of 790 patients for whom data from two time-points was available, yielding 91 (13%) who showed severe depressive symptoms at both time-points. While training a baseline model on the study population inpatients #1 dataset alone and testing its generalization to the nine other sites was not feasible as study population inpatients #1 contained only 19 (14%) patients who were severely depressed at both time-points, we assessed our ability to predict severe depression without treatment response using leave-site-out cross-validation. In this procedure, data from all but one site is used for training and the model is tested on the remaining site. This is repeated for each site. To counter the strong class imbalance, we employed the elastic net approach for classification with Synthetic Minority Over-sampling Technique (SMOTE) combined with Edited Nearest Neighbors (ENN) as proposed by Batista et. al (*58*). With this approach, we show that presence/absence of persistent, severe depression can be predicted with an average balanced accuracy of .66. Performance per site ranges from chance level (balanced accuracy = .50) in the real-world outpatients #6 dataset to .86 in study population inpatients #2 (Table S3).

Supplementary  analysis of generalization performance excluding neuroticism

To determine the extent to which the generalizability of our predictive model relied on neuroticism, a trait strongly correlated with depressive symptoms (*59*), we conducted an additional analysis to explicitly test the generalization performance of our model when excluding neuroticism. To this end, we retrained the machine learning model on the remaining top 4 predictive features on study population #1 and tested its performance on all other sites. As expected, generalization performance across all external datasets decreased from r(2,673)=.60 to r(2,673)=.50, but remained significant (p<.001) when excluding Neuroticism. Likewise, Standard Deviation increased from .089 to .121. Investigating performance on the nine samples separately excluding Neuroticism shows that performance on all sites decreases to r(1,227)=.26 in the real-world general population sample, remains unchanged in real-world outpatients #6 (r(250)=.50), and only slightly decreases to r(350)=.69 in real-world inpatients #1 (from r(350)=.73). Thus, all but the lowest performances (real-world general population sample) lie within 1.41 standard deviations of the mean of the base model performance without Neuroticism (r(364)=.62, standard deviation=.123) indicating good generalization even without the most highly weighted feature of the original model.

**Fig. S1. Feature Permutation Importance from Elastic Net Modeling.**
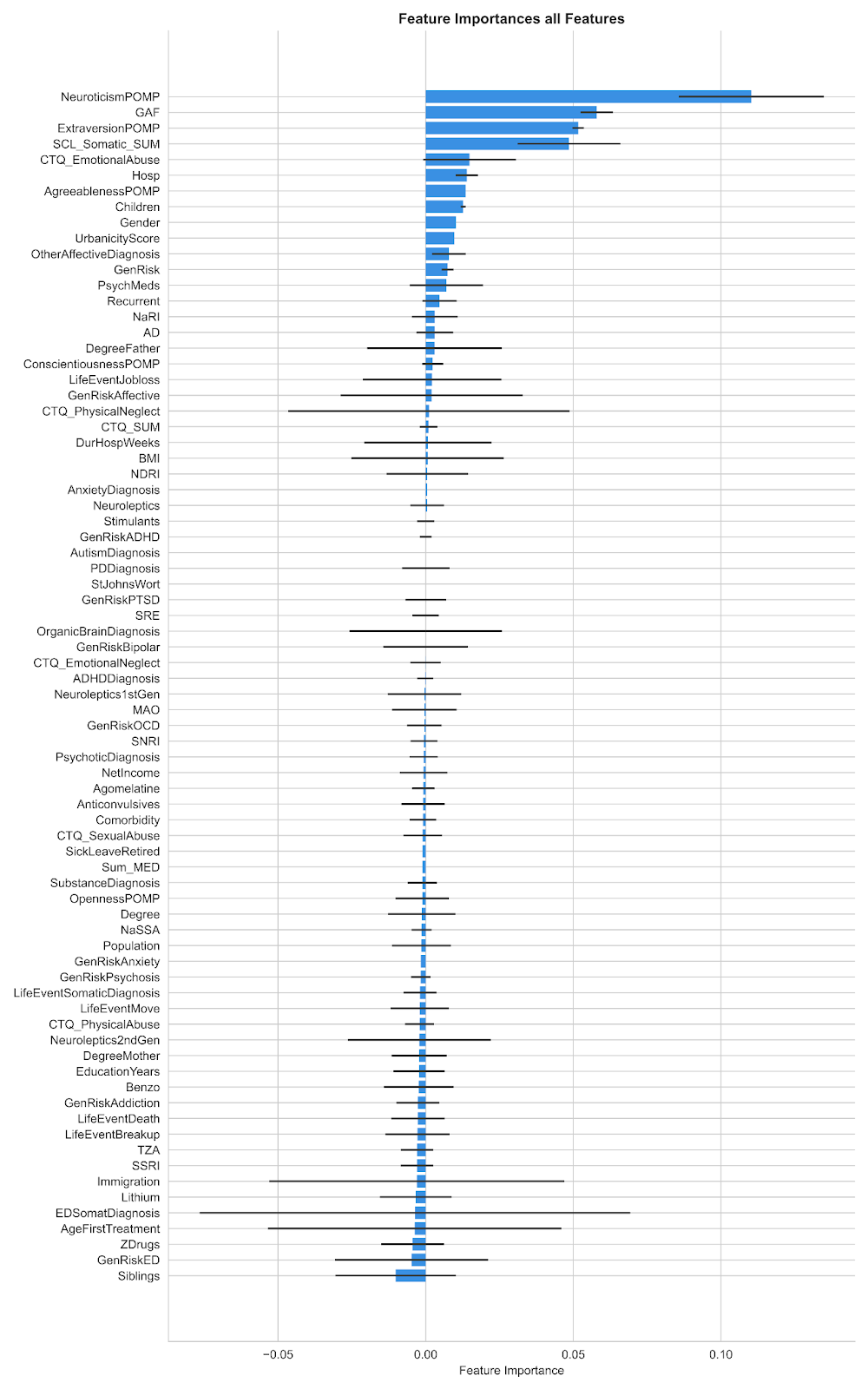

Permutation importance ranking of features is depicted, gauging the influence of each on the model's predictive error when permuted. The vertical bars manifest the average importance scores, and the error bars extending horizontally denote one standard deviation from this mean, capturing the consistency of feature significance. This representation underscores the relative contribution and stability of each feature within the model's architecture.

**Fig. S2. Mean Absolute Error (MAE) Analysis for Base Model Predictions.**

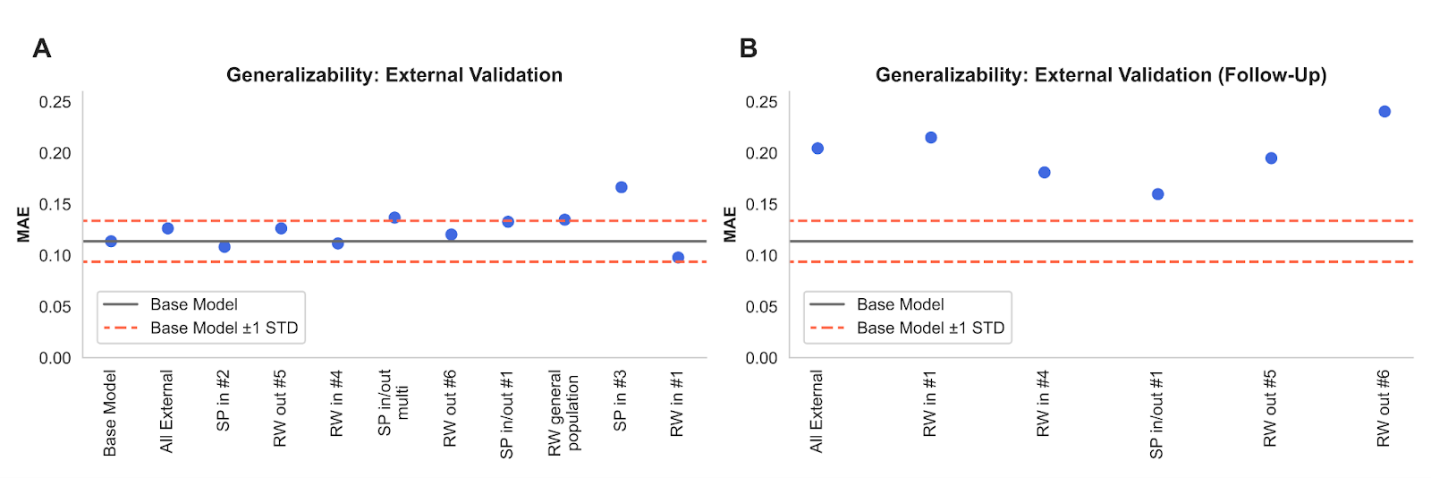

(A) Baseline Depression Severity Predictions: Model performance across nine external validation sites, with a solid line indicating the base model's MAE and dashed lines representing the MAE ±1 STD. Individual data points denote the site-specific MAE. (B) Follow-Up Depression Severity Predictions: Comparable depiction of the model’s predictive accuracy for follow-up depression severity scores.

**Table S1.**

Somatic comorbidities that served as exclusion criteria for study population inpatients, site #1.

| **Automatic exclusion from participation** | **Inclusion only on case by case basis, depending on medication status** |
| --- | --- |
| Ankylosing spondylitis with HLA B27 antigen | Allergies |
| Carcinoma | Asthma |
| Colitis Ulcerosa | Borreliosis |
| Crohn’s disease | Chronic bronchitis |
| Encephalopathy | Chronic leukozytosis |
| Epilepsy | Diabetes mellitus type I |
| Hereditary postural tremor | Hemochromatosis |
| Facioscapulohumeral muscular dystrophy | Hashimoto’s thyreoidits |
| Hepatitis C | Hepatitis B |
| Lupus | Lichen sclerosus |
| Multiple Sclerosis | Lymphocytic thyroiditis |
| Narcolepsy | Meningitis |
| Neuroborreliosis | Migraine with aura |
| Neural muscle atrophy | Atopic dermatitis |
| Sarcoidosis | Neurofibromatosis type I |
| Sjögren syndrome | Paroxysmal hemicrania |
| Scleroderma | Proctitis ulcerosa |
| Undifferenciated connective tissue disease | Rheumatic disorders |
| Stroke | Restless legs syndrome |
| Malignant or semi-malignant tumor | Sleep apnea |
| Tumor in the head region (including benign) | Tinnitus |
| Rheumatoid arthritis | Tuberculosis |
|  | Gitelman syndrome |
|  | Psoriasis |
|  | Disorders of antidiuretic hormone (ADH) secretion |

**Table S2.**

Group comparisons between study population and real-world inpatients from site #1.

| **Dimension** | **Variables** | **Study population inpatients, site #1** | **N** | **Real-world inpatients, site #1** | **N** | **Test statistic** | **p** | **p_FDR_** |
| --- | --- | --- | --- | --- | --- | --- | --- | --- |
| **Dimension 1: Sociodemographic variables** |  |  |  |  |  |  |  |  |
|  | **Age** |  |  |  |  |  |  |  |
|  | Mean  (SD) | 37.48  (13.10) | 388 | 39.65  (17.27) | 360 | -1.92 ^b^ | .055 | .112 |
|  | Range | 18-66 |  | 18-81 |  |  |  |  |
|  | **Gender** |  |  |  |  |  |  |  |
|  | male/female | 166/222 | 388 | 170/190 | 360 | 1.49 ^b^ | .239 | .344 |
|  | **Years in formal education** |  |  |  |  |  |  |  |
|  | Mean  (SD) | 13.83  (2.56) | 373 | 12.63  (2.79) | 360 | 6.06 ^b^ | **<.001** | **.003** |
|  | **Highest degree (self)** |  |  |  |  |  |  |  |
|  | Mean rank | 414.48 | 369 | 304.94 | 352 | 45210.5 ^c^ | **<.001** | **.003** |
|  | **Highest degree (father)** |  |  |  |  |  |  |  |
|  | Mean rank | 281.93 | 220 | 285.31 | 347 | 37715.0 ^c^ | .809 | .841 |
|  | **Highest degree (mother)** |  |  |  |  |  |  |  |
|  | Mean rank | 276.83 | 222 | 289.42 | 346 | 36702.5.5 ^c^ | .365 | .472 |
|  | **Household income** |  |  |  |  |  |  |  |
|  | Mean  (SD) | 2557.94  (1899.40) | 188 | 2528.37 (4791.98) | 320 | .081 ^b^ | .936 | .945 |
|  | **Population birth place** |  |  |  |  |  |  |  |
|  | Mean  (SD) | 137185.92  (281887.78) | 154 | 221352.41  (1292214.69) | 343 | -0.799 ^b^ | .424 | .533 |
|  | **Urbanicity Score** |  |  |  |  |  |  |  |
|  | Mean Rank | 277.32 | 145 | 230.63 | 343 | 20108.5 ^c^ | **<.001** | **.003** |
|  | **Immigration** |  |  |  |  |  |  |  |
|  | yes/no | 27/180 | 207 | 63/289 | 352 | 2.27 ^b^ | .153 | .250 |
|  | **Children** |  |  |  |  |  |  |  |
|  | yes/no | 73/155 | 228 | 138/214 | 352 | 3.09 ^b^ | .093 | .179 |
|  | **Siblings** |  |  |  |  |  |  |  |
|  | yes/no | 206/21 | 227 | 316/36 | 352 | .148 ^b^ | .776 | .831 |
| **Dimension 2: Depressive symptom**  **severity** |  |  |  |  |  |  |  |  |
|  | **BDI** |  |  |  |  |  |  |  |
|  | Mean  (SD) | 25.08  (10.72) | 366 | 24.76  (11.40) | 352 | .389 ^b^ | .698 | .771 |
|  | **HAMD** |  |  |  |  |  |  |  |
|  | Mean  (SD) | 15.85  (6.47) | 385 | 17.13  (6.54) | 329 | -2.61 ^b^ | **.009** | **.025** |
|  | **GAF** |  |  |  |  |  |  |  |
|  | Mean  (SD) | 54.25  (9.47) | 385 | 53.73  (9.36) | 341 | .748 ^b^ | .454 | .554 |
|  | **Recurring depression** |  |  |  |  |  |  |  |
|  | yes/no | 289/98 | 387 | 238/122 | 360 | 6.59 ^b^ | **.013** | **.034** |
|  | **Psychiatric comorbidity** |  |  |  |  |  |  |  |
|  | yes/no | 219/168 | 387 | 164/196 | 360 | 9.09 ^b^ | **.003** | **.010** |
| **Dimension 3: Current psychotropic medication** |  |  |  |  |  |  |  |  |
|  | **Medication load** |  |  |  |  |  |  |  |
|  | Mean  (SD) | 2.61  (1.83) | 388 | 2.51  (2.22) | 296 | .638 ^b^ | .524 | .617 |
|  | **Psychotropic medication** |  |  |  |  |  |  |  |
|  | yes/no | 352/36 | 388 | 305/55 | 360 | 6.29 ^a^ | **.014** | **.035** |
|  | **Antidepressant** |  |  |  |  |  |  |  |
|  | yes/no | 335/53 | 388 | 296/91 | 360 | 16.21 ^a^ | **<.001** | **.003** |
|  | **Agomelatine** |  |  |  |  |  |  |  |
|  | yes/no | 40/348 | 388 | 20/340 | 360 | 5.72 ^a^ | **.021** | **.048** |
|  | **TCA** |  |  |  |  |  |  |  |
|  | yes/no | 28/360 | 388 | 22/338 | 360 | .366 ^a^ | .562 | .648 |
|  | **NRI** |  |  |  |  |  |  |  |
|  | yes/no | 2/386 | 388 | 1/359 | 360 | .264 ^a^ | 1.0 | .999 |
|  | **NDRI** |  |  |  |  |  |  |  |
|  | yes/no | 22/366 | 388 | 29/331 | 360 | 1.67 ^a^ | .245 | .344 |
|  | **SSRI** |  |  |  |  |  |  |  |
|  | yes/no | 97/291 | 388 | 99/261 | 360 | .604^a^ | .455 | .554 |
|  | **SNRI** |  |  |  |  |  |  |  |
|  | yes/no | 186/202 | 388 | 125/235 | 360 | 13.43 ^a^ | **<.001** | **.003** |
|  | **NaSSA** |  |  |  |  |  |  |  |
|  | yes/no | 92/296 | 388 | 91/269 | 360 | .248 ^a^ | .671 | .749 |
|  | **MAOI** |  |  |  |  |  |  |  |
|  | yes/no | 8/380 | 388 | 6/354 | 360 | .159 ^a^ | .791 | .831 |
|  | **SRE** |  |  |  |  |  |  |  |
|  | yes/no | 0/388 | 388 | 2/358 | 360 | 2.16 ^a^ | .231 | .340 |
|  | **Neuroleptics** |  |  |  |  |  |  |  |
|  | yes/no | 162/226 | 388 | 172/188 | 360 | 2.74 ^a^ | .106 | .194 |
|  | **1^st^ generation neuroleptics** |  |  |  |  |  |  |  |
|  | yes/no | 42/342 | 388 | 52/308 | 360 | 1.10 ^a^ | .329 | .431 |
|  | **2^nd^ generation neuroleptics** |  |  |  |  |  |  |  |
|  | yes/no | 140/248 | 388 | 147/213 | 360 | 1.78 ^a^ | .201 | .304 |
|  | **Lithium** |  |  |  |  |  |  |  |
|  | yes/no | 20/368 | 388 | 11/349 | 360 | 2.07 ^a^ | .198 | .304 |
|  | **Anticonvulsives** |  |  |  |  |  |  |  |
|  | yes/no | 11/377 | 388 | 19/341 | 360 | 2.89 ^a^ | .096 | .182 |
|  | **Stimulants** |  |  |  |  |  |  |  |
|  | yes/no | 1/387 | 388 | 9/351 | 360 | 7.12 ^a^ | **.009** | **.025** |
|  | **Benzodiazepines** |  |  |  |  |  |  |  |
|  | yes/no | 4/384 | 388 | 44/316 | 360 | 38.95 ^a^ | **<.001** | **.003** |
|  | **Z-drugs** |  |  |  |  |  |  |  |
|  | yes/no | 6/382 | 388 | 26/334 | 360 | 14.69 ^a^ | **<.001** | **.003** |
| **Dimension 4: Family and own**  **psychiatric history** |  |  |  |  |  |  |  |  |
|  | **Genetic risk total** |  |  |  |  |  |  |  |
|  | yes/no | 117/86 | 203 | 177/142 | 319 | .233 ^a^ | .652 | .736 |
|  | **Genetic risk major depressive disorder** |  |  |  |  |  |  |  |
|  | yes/no | 74/152 | 226 | 125/227 | 352 | .467 ^a^ | .530 | .617 |
|  | **Genetic risk bipolar disorder** |  |  |  |  |  |  |  |
|  | yes/no | 1/214 | 215 | 16/336 | 360 | 7.64 ^a^ | **.004** | **.012** |
|  | **Genetic risk substance use disorder** |  |  |  |  |  |  |  |
|  | yes/no | 29/194 | 223 | 50/302 | 352 | .166 ^a^ | .711 | .777 |
|  | **Genetic risk anxiety disorder** |  |  |  |  |  |  |  |
|  | yes/no | 9/206 | 215 | 27/325 | 352 | 2.73 ^a^ | .112 | .198 |
|  | **Genetic risk obsessive-compulsive disorder** |  |  |  |  |  |  |  |
|  | yes/no | 3/212 | 215 | 4/348 | 352 | .073 ^a^ | 1.0 | .999 |
|  | **Genetic risk posttraumatic stress disorder** |  |  |  |  |  |  |  |
|  | yes/no | 1/214 | 215 | 14/338 | 352 | 6.39 ^a^ | **.013** | **.034** |
|  | **Genetic risk psychosis** |  |  |  |  |  |  |  |
|  | yes/no | 9/207 | 216 | 12/340 | 352 | .216 ^a^ | .653 | .736 |
|  | **Genetic risk ADHD** |  |  |  |  |  |  |  |
|  | yes/no | 1/214 | 215 | 13/339 | 352 | 5.78 ^a^ | **.022** | **.049** |
|  | **Genetic risk eating disorder** |  |  |  |  |  |  |  |
|  | yes/no | 5/209 | 214 | 17/335 | 352 | 2.21 ^a^ | .179 | .287 |
|  | **Age at first psychiatric treatment** |  |  |  |  |  |  |  |
|  | Mean  (SD) | 31.49  (12.47) | 356 | 28.84  (15.99) | 348 | 2.45 ^b^ | **.015** | **.036** |
|  | **Number of hospitalizations** |  |  |  |  |  |  |  |
|  | Mean  (SD) | 2.31  (2.02) | 386 | 2.86  (4.67) | 349 | -2.05 ^b^ | **.041** | .089 |
|  | **Duration of hospitalizations**  **(total in weeks)** |  |  |  |  |  |  |  |
|  | Mean  (SD) | 10.60  (17.70) | 384 | 31.63  (34.91) | 243 | -8.71 ^b^ | **<.001** | **.003** |
|  | **Duration on sick leave/retired (due to mental illness)**  **(total in months)** |  |  |  |  |  |  |  |
|  | Mean  (SD) | 10.62  (31.18) | 349 | 19.18  (45.70) | 348 | -2.89 ^b^ | **.004** | **.012** |
| **Dimension 5: Childhood maltreatment and stressful life events** |  |  |  |  |  |  |  |  |
|  | **CTQ Sum** |  |  |  |  |  |  |  |
|  | Mean  (SD) | 46.33  (16.17) | 362 | 47.17  (16.90) | 345 | -.673 ^b^ | .501 | .603 |
|  | **CTQ physical neglect** |  |  |  |  |  |  |  |
|  | Mean  (SD) | 8.10  (3.01) | 365 | 11.55  (5.82) | 350 | -9.89 ^b^ | **<.001** | **.003** |
|  | **CTQ emotional neglect** |  |  |  |  |  |  |  |
|  | Mean  (SD) | 13.87  (5.62) | 366 | 13.76  (5.79) | 348 | .264 ^b^ | .792 | .831 |
|  | **CTQ physical abuse** |  |  |  |  |  |  |  |
|  | Mean  (SD) | 7.03  (3.52) | 365 | 7.09  (3.66) | 349 | -.230 ^b^ | .718 | .777 |
|  | **CTQ emotional abuse** |  |  |  |  |  |  |  |
|  | Mean  (SD) | 11.19  (5.49) | 364 | 11.55  (5.82) | 350 | -.855 ^b^ | .393 | .502 |
|  | **CTQ sexual abuse** |  |  |  |  |  |  |  |
|  | Mean  (SD) | 6.25  (3.37) | 365 | 6.69  (3.75) | 348 | -1.63 ^b^ | .105 | .194 |
|  | **Separation or divorce** |  |  |  |  |  |  |  |
|  | yes/no | 37/124 | 161 | 62/289 | 351 | 2.00 ^a^ | .185 | .290 |
|  | **Death of a loved one** |  |  |  |  |  |  |  |
|  | yes/no | 21/140 | 161 | 57/294 | 351 | .873 ^a^ | .427 | .533 |
|  | **Job loss** |  |  |  |  |  |  |  |
|  | yes/no | 24/137 | 161 | 39/312 | 351 | 1.47 ^a^ | .247 | .344 |
|  | **Moved house** |  |  |  |  |  |  |  |
|  | yes/no | 31/130 | 161 | 70/281 | 351 | .033 ^a^ | .905 | .922 |
| **Dimension 6: Somatic symptoms** |  |  |  |  |  |  |  |  |
|  | **SCL-90-R somatization subscale** |  |  |  |  |  |  |  |
|  | Mean  (SD) | 10.88  (8.43) | 199 | 12.25 (7.80) | 353 | -1.92 ^b^ | .055 | .112 |
|  | **BMI** |  |  |  |  |  |  |  |
|  | Mean  (SD) | 26.01  (5.44) | 348 | 27.33  (8.76) | 46 | -.996 ^b^ | .324 | .429 |
| **Dimension 7: Big Five Personality Dimensions** |  |  |  |  |  |  |  |  |
|  | **Neuroticism** |  |  |  |  |  |  |  |
|  | Mean  (SD) | 66.84  (15.59) | 352 | 68.70  (17.39) | 345 | -1.48 ^b^ | .138 | .240 |
|  | **Extraversion** |  |  |  |  |  |  |  |
|  | Mean  (SD) | 39.24  (14.57) | 354 | 36.13  (18.65) | 343 | 2.45 ^b^ | **.015** | **.036** |
|  | **Openness** |  |  |  |  |  |  |  |
|  | Mean  (SD) | 58.10  (14.05) | 351 | 55.03  (20.64) | 344 | 2.29 ^b^ | **.023** | .051 |
|  | **Agreeableness** |  |  |  |  |  |  |  |
|  | Mean  (SD) | 64.55  (13.18) | 351 | 68.16  (15.88) | 346 | -3.26 ^b^ | **.001** | **.003** |
|  | **Conscientiousness** |  |  |  |  |  |  |  |
|  | Mean  (SD) | 60.42  (15.79) | 352 | 55.74  (19.09) | 346 | 3.52 ^b^ | **<.001** | **.003** |

*Abbreviations.* ADHD = Attention Deficit Hyperactivity Disorder, BDI = Beck Depression Inventory, BMI = Body Mass Index, CTQ = Childhood Maltreatment Questionnaire, GAF = General Assessment of Functioning, HAMD = Hamilton Depression Rating Scale, MAOI = Monoamine Oxidase Inhibitors, NaSSA = Noradrenergic and Specific Serotonergic Antidepressants, NDRI = Norepinephrine–Dopamine Reuptake Inhibitor , NRI = Norepinephrine Reuptake Inhibitor, SCL = Symptom Checklist, SNRI = Serotonin–Norepinephrine Reuptake Inhibitor, SRE = Serotonin Reuptake Enhancer, SSRI = Selective Serotonin Reuptake Inhibitor, TCA = Tricyclic Antidepressant.

*Note.* Analysis method is indicated as follows: ^a^ X²-test, ^b^ two-sample t-test, ^c^ Mann-Whitney U test.

**Table S3. Results for classification of severely depressed non-responders.**

| **Site** | **N_train** | **N_test** | **balanced accuracy** |
| --- | --- | --- | --- |
| Study population inpatients, site #2 | 723 | 67 | 0.860656 |
| Real-world inpatients, site #4 | 662 | 128 | 0.713705 |
| Study population in-& outpatients, site #1 | 732 | 58 | 0.680000 |
| Study population inpatients, site #1 | 635 | 155 | 0.652864 |
| Real-world inpatients, site #1 | 617 | 173 | 0.634588 |
| Real-world outpatients, site #5 | 708 | 82 | 0.580128 |
| Real-world outpatients, site #6 | 663 | 127 | 0.500000 |

56.	A. Peters, K. H. Greiser, S. Göttlicher, W. Ahrens, M. Albrecht, F. Bamberg, T. Bärnighausen, H. Becher, K. Berger, A. Beule, H. Boeing, B. Bohn, K. Bohnert, B. Braun, H. Brenner, R. Bülow, S. Castell, A. Damms-Machado, M. Dörr, N. Ebert, M. Ecker, C. Emmel, B. Fischer, C. W. Franzke, S. Gastell, G. Giani, M. Günther, K. Günther, K. P. Günther, J. Haerting, U. Haug, I. M. Heid, M. Heier, D. Heinemeyer, T. Hendel, F. Herbolsheimer, J. Hirsch, W. Hoffmann, B. Holleczek, H. Hölling, A. Hörlein, K. H. Jöckel, R. Kaaks, A. Karch, S. Karrasch, N. Kartschmit, H. U. Kauczor, T. Keil, Y. Kemmling, B. Klee, B. Klüppelholz, A. Kluttig, L. Kofink, A. Köttgen, D. Kraft, G. Krause, L. Kretz, L. Krist, J. Kühnisch, O. Kuß, N. Legath, A. T. Lehnich, M. Leitzmann, W. Lieb, J. Linseisen, M. Loeffler, A. Macdonald, K. H. Maier-Hein, N. Mangold, C. Meinke-Franze, C. Meisinger, J. Melzer, B. Mergarten, K. B. Michels, R. Mikolajczyk, S. Moebus, U. Mueller, M. Nauck, T. Niendorf, K. Nikolaou, N. Obi, S. Ostrzinski, L. Panreck, I. Pigeot, T. Pischon, I. Pschibul-Thamm, W. Rathmann, A. Reineke, S. Roloff, D. Rujescu, S. Rupf, O. Sander, T. Schikowski, S. Schipf, P. Schirmacher, C. L. Schlett, B. Schmidt, G. Schmidt, M. Schmidt, G. Schöne, H. Schulz, M. B. Schulze, A. Schweig, A. M. Sedlmeier, S. Selder, J. Six-Merker, R. Sowade, A. Stang, O. Stegle, K. Steindorf, G. Stübs, E. Swart, H. Teismann, I. Thiele, S. Thierry, M. Ueffing, H. Völzke, S. Waniek, A. Weber, N. Werner, H. E. Wichmann, S. N. Willich, K. Wirkner, K. Wolf, R. Wolff, H. Zeeb, M. Zinkhan, J. Zschocke, Framework and baseline examination of the German National Cohort (NAKO). *Eur J Epidemiol* **37**, 1107–1124 (2022).

57.	R. Rosenthal, D. B. Rubin, A simple, general purpose display of magnitude of experimental effect. *J Educ Psychol* **74**, 166 (1982).

58.	G. E. Batista, R. C. Prati, M. C. Monard, A study of the behavior of several methods for balancing machine learning training data. *ACM SIGKDD explorations newsletter* **6**, 20–29 (2004).

59.	L. B. Navrady, S. J. Ritchie, S. W. Y. Chan, D. M. Kerr, M. J. Adams, E. H. Hawkins, D. Porteous, I. J. Deary, C. R. Gale, G. D. Batty, A. M. McIntosh, Intelligence and neuroticism in relation to depression and psychological distress: Evidence from two large population cohorts. *European Psychiatry* **43**, 58–65 (2017).
